# Prevalence and determinants of oral and cervicogenital HPV infection: baseline analysis of the MHOC cohort study

**DOI:** 10.1101/2022.01.15.22269344

**Authors:** Andrew F. Brouwer, Lora P. Campredon, Heather M. Walline, Brittany M. Marinelli, Christine M. Goudsmit, Trey B. Thomas, Rachel L. Delinger, Yan Kwan Lau, Emily C. Andrus, Monica L. Yost, Jodi K. McCloskey, Taylor S. Sullivan, Alex S. Mortensen, Suiyuan Huang, Keith Murphy, Bonnie Cheng, Kayla Stanek, Thankam Nair, Thomas E. Carey, Rafael Meza, Marisa C. Eisenberg

## Abstract

We determined baseline oral and cervicogenital human papillomavirus (HPV) prevalence and determinants of infection in the Michigan HPV and Oropharyngeal Cancer (MHOC) study. We enrolled 394 college-age and older-adult participants of both sexes in Ann Arbor, Michigan and the surrounding area. All participants provided an oral sample at baseline, and 130 females provided a cervicogenital sample. Samples were tested for 18 HPV genotypes using polymerase chain reaction (PCR) MassArray. Participants filled out sociodemographic and behavioral questionnaires. Prevalence ratios for HPV oral or cervicogenital prevalence by predictor variables were estimated in univariable log-binomial models. Analysis was conducted 2018–20. In the full cohort, baseline oral HPV prevalence was 10.0% for any detected genotype (among the 338 valid oral tests at baseline) and 6.5% for high-risk types, and cervicogenital prevalence was 20.0% and 10.8%, respectively (among the 130 first valid cervicogenital tests). Oral HPV prevalence did not vary by sex, with 10.5% of women and 9.0% of men having an infection. We found a high prevalence of oral and cervicogenital HPV infection among those reporting no recent sexual partners compared to those with a single recent sexual partner, but prevalence increased with the number of recent partners for most sexual behaviors. We observed an ecological fallacy masking the direction of impact of vaccination on HPV prevalence in the full cohort compared to the college-aged and older-adult populations considered separately. Substance use was not significantly associated with oral or cervicogenital HPV infection. Many studies report substantially higher oral HPV infection prevalence in men than in women. That difference may not be uniform across populations in the US.

## Introduction

While the human papillomavirus (HPV) is well known as the primary cause of cervical cancer, incidence of HPV-related oropharyngeal squamous cell carcinoma (OPSCC) in the U.S. is now greater than that of cervical cancer [1–4]. Indeed, an increasing fraction of OPSCCs are attributable to HPV [5–8], with some studies reporting over 80% of OPSCC cancer patients testing positive for high-risk HPV [9]. Understanding the determinants, prevalence, and dynamics of oral HPV will be essential for designing interventions.

OPSCC is more than three times more common in men than in women [1, 4], which mirrors higher prevalence of oral HPV in men. Nationally representative studies in the U.S. have estimated prevalence of oral HPV as approximately 11% in men and 4% in women [10, 11]. Oral sex is a known risk factor—and likely primary transmission pathway—for oral HPV [12–15], and there may be risk differences based on the sex of one’s partner [16]. More broadly, sexual behaviors and smoking have been identified as important risk factors of oral HPV infection [17]. Nevertheless, questions remain about which risk factors are in fact causally related to transmission and which are correlated with transmission behaviors. Moreover, the impact that HPV vaccination will have on oral HPV prevalence [18, 19] and transmission dynamics [20] needs to be elucidated.

The Michigan HPV and Oropharyngeal Cancer (MHOC) Study aims to evaluate patterns of oral HPV infection, prevalence, incidence and clearance and their relationship to sexual history and sexual behaviors. MHOC includes an epidemiological arm that tests a longitudinal cohort for oral (and, in a subset of participants, cervicogential) HPV [21]. The longitudinal HPV infection and sexual history data will facilitate the development of individual-based network models of HPV transmission and will be used to parameterize multiscale models of HPV-related OPSC carcinogenesis. This first, hypothesis-generating analysis, we present the baseline prevalence of oral and cervicogenital HPV and the association of infection with demographic and behavioral characteristics in the MHOC Study.

## Methods

We previously published the full study protocol [21], including both the baseline and longitudinal aspects of the study. Here, we briefly describe the main aspects of the study related to the baseline prevalence examined here.

### Study subjects

Study participants were recruited in Ann Arbor, Michigan and the immediate surrounding areas. Participants were recruited at University of Michigan campus residence halls, through community fliers, and through the UM Health Research website. Volunteers over the age of 18 without a history of head and neck cancer who were willing to participate in both the baseline and longitudinal portions of the study were invited to enroll. The full inclusion/exclusion criteria are given in [21]. We enrolled 394 participants between April 2015 and December 2017. A substudy focusing on cervicogenital HPV enrolled 130 participants. Documented informed consent was obtained from all participants. The University of Michigan IRB approved consent documents and study protocol (HUM00090236).

### Surveys

A baseline questionnaire was administered to each participant at their initial visit. Participant ID numbers were assigned to ensure participant confidentiality. The surveys were designed to individually assess a variety of topics including demographics, vaccination and screening history, sexual health and behavior, and alcohol and drug use. Sexual behavior questions assessed current and past experiences of vaginal, oral, and anal sex.

### HPV testing

All participants self-collected a saliva sample with Scope mouthwash (Proctor & Gamble; Cincinnati, OH) or an Oragene RE-100 kit (DNA Genotek; Kanata, Canada). Saliva samples were taken at each study visit. Participants who had a vagina, were not pregnant, and were not menstruating at the time of a study visit were invited to self-collect a cervicovaginal sample with a HerSwab (Eve Medical; Toronto, Canada). DNA was extracted from samples and genotyped using PCR MassArray [9]. We tested for 18 genotypes: 6, 11, 16, 18, 31, 33, 35, 39, 45, 51, 52, 56, 58, 59, 66, 68, 73, and 90. More details on the testing are given in [9, 21]. We further categorized genotypes by those included in the cobas^®^ HPV test (Roche Diagnostics; Risch-Rotkreuz, Switzerland) used in clinical settings, those designated by the International Agency for Research on Cancer (IARC) as group 1 (carcinogenic) or group 2A (probably carcinogenic) carcinogens [22], those included in the Gardasil and Gardasil 9 vaccines (Merck; Kenilworth, NJ), and those responsible for genital warts.

### Statistical analysis

Study data were collected and managed using REDCap electronic data capture tools hosted at the University of Michigan [23, 24]. Data were analyzed in R 3.6 [25], 2018–20.

We consider two main outcome variables: 1) presence of HPV in a participant’s oral sample at the baseline study visit and 2) presence of HPV in a participant’s first valid cervicogenital sample. We use baseline oral and first valid cervicogenital test because the cervicogenital substudy was rolled out after the main study began. We assessed sample prevalence of these two outcomes in their respective populations based on demographic and behavior variables derived from participant answers to the surveys. We also stratified the cohort into ages 18–22 years (college-age cohort) and ages 23+ (older cohort) for oral HPV prevalence. To estimate prevalence ratios, we ran univariable, log-binomial regression models for oral HPV prevalence predicted by the variables in Table 1 in the full cohort and for cervicogenital HPV prevalence in the substudy cohort. We also separately modeled oral HPV prevalence in the college-age and older cohorts. Participants with missing data for a predictor variable were excluded from that model.

**Table 1:** Baseline characteristics of participants in the MHOC Study (data collected in Ann Arbor, MI, 2015-17, analyzed 2018-2020). Note: percentages may not add up to 100% as participants could refuse to answer questions. *Other than HPV. †Number of partners in the past 6 months among those who ever engaged in the behavior.

|  | Full cohort<br>(N=394) |  | College-age<br>cohort<br>(N= 241) |  | Older adult<br>cohort<br>(N=153) |  | Cervicogenital<br>substudy cohort<br>(N= 130) |  |
| --- | --- | --- | --- | --- | --- | --- | --- | --- |
|  | % | n | % | n | % | n | % | n |
| **Age (years)** |  |  |  |  |  |  |  |  |
| 18 | 25% | 99 | 41% | 99 | 0% | 0 | 27% | 35 |
| 19-22 | 36% | 142 | 59% | 142 | 0% | 0 | 33% | 43 |
| 23-29 | 12% | 49 | 0% | 0 | 32% | 49 | 11% | 14 |
| 30-49 | 12% | 46 | 0% | 0 | 30% | 46 | 14% | 19 |
| 50+ | 15% | 58 | 0% | 0 | 38% | 58 | 15% | 19 |
| **Sex** |  |  |  |  |  |  |  |  |
| Female | 67% | 265 | 70% | 168 | 63% | 97 | 100% | 130 |
| Male | 33% | 129 | 30% | 73 | 37% | 56 | 0% | 0 |
| **Race** |  |  |  |  |  |  |  |  |
| White | 61% | 240 | 54% | 131 | 71% | 109 | 62% | 80 |
| Asian | 21% | 83 | 31% | 74 | 6% | 9 | 20% | 26 |
| Black/Hispanic/multiracial/unknown | 18% | 71 | 15% | 36 | 23% | 35 | 18% | 24 |
| **Marital status** |  |  |  |  |  |  |  |  |
| Never married | 77% | 302 | 98% | 237 | 42% | 65 | 75% | 97 |
| Married/partnered | 19% | 73 | 1% | 2 | 46% | 71 | 19% | 25 |
| Widowed/divorced/separated | 4% | 17 | 0% | 1 | 10% | 16 | 6% | 8 |
| **Sexual attraction** |  |  |  |  |  |  |  |  |
| Mostly to only to opposite sex | 89% | 41 | 87% | 210 | 87% | 133 | 92% | 120 |
| Equal to only to same sex | 10% | 343 | 10% | 25 | 10% | 16 | 5% | 6 |
| **Circumcised (male only)** |  |  |  |  |  |  |  |  |
| Yes | 71% | 91 | 66% | 48 | 77% | 43 | --- | --- |
| No | 29% | 37 | 33% | 24 | 23% | 13 | --- | --- |
| **Ever diagnosed with STI\*** |  |  |  |  |  |  |  |  |
| No | 94% | 369 | 98% | 237 | 86% | 132 | 93% | 121 |
| Yes | 6% | 25 | 2% | 4 | 14% | 21 | 7% | 9 |
| **HPV vaccination** |  |  |  |  |  |  |  |  |
| No | 44% | 177 | 24% | 59 | 77% | 118 | 45% | 58 |
| Yes | 49% | 192 | 66% | 158 | 22% | 34 | 51% | 66 |
| **Alcohol use** |  |  |  |  |  |  |  |  |
| Never or non-current | 29% | 115 | 31% | 75 | 26% | 40 | 28% | 37 |
| Current | 69% | 260 | 65% | 157 | 74% | 113 | 70% | 91 |
| **Ever cigarette use** |  |  |  |  |  |  |  |  |
| Never | 76% | 300 | 87% | 209 | 59% | 91 | 79% | 103 |
| Ever | 23% | 91 | 12% | 30 | 40% | 61 | 20% | 26 |
| **Ever marijuana use** |  |  |  |  |  |  |  |  |
| Never | 52% | 204 | 59% | 141 | 41% | 63 | 52% | 68 |
| Ever | 43% | 171 | 36% | 87 | 55% | 84 | 45% | 59 |
| **Has ever engaged in** |  |  |  |  |  |  |  |  |
| Deep kissing |  |  |  |  |  |  |  |  |
| Yes | 84% | 330 | 80% | 194 | 89% | 136 | 88% | 115 |
| No | 16% | 62 | 19% | 46 | 10% | 16 | 12% | 15 |
| Manual sex |  |  |  |  |  |  |  |  |
| Yes | 68% | 268 | 60% | 144 | 81% | 124 | 73% | 95 |
| No | 29% | 116 | 38% | 92 | 16% | 24 | 25% | 33 |
| Vaginal, oral, or anal sex |  |  |  |  |  |  |  |  |
| Yes | 75% | 296 | 63% | 152 | 94% | 144 | 78% | 101 |
| No | 24% | 95 | 36% | 86 | 6% | 9 | 22% | 29 |
| Vaginal sex |  |  |  |  |  |  |  |  |
| Yes | 64% | 252 | 48% | 116 | 89% | 136 | 68% | 89 |
| No | 35% | 137 | 51% | 122 | 10% | 15 | 32% | 41 |
| Oral sex |  |  |  |  |  |  |  |  |
| Yes | 71% | 279 | 60% | 145 | 88% | 134 | 75% | 98 |
| No | 28% | 110 | 39% | 93 | 11% | 17 | 25% | 32 |
| Anal sex |  |  |  |  |  |  |  |  |
| Yes | 22% | 88 | 12% | 30 | 38% | 58 | 20% | 26 |
| No | 76% | 299 | 86% | 208 | 60% | 91 | 78% | 102 |
| Anilingus |  |  |  |  |  |  |  |  |
| Yes | 13% | 50 | 7% | 17 | 22% | 33 | 10% | 13 |
| No | 86% | 340 | 92% | 222 | 77% | 118 | 90% | 117 |
| **Deep kissing partners†** |  |  |  |  |  |  |  |  |
| 0 | 13% | 51 | 8% | 20 | 20% | 31 | 14% | 18 |
| 1 | 42% | 165 | 33% | 80 | 56% | 85 | 42% | 54 |
| 2-4 | 20% | 77 | 24% | 59 | 12% | 18 | 24% | 31 |
| 5+ | 9% | 37 | 15% | 35 | 1% | 2 | 9% | 12 |
| **Vaginal, oral, or anal sex partners†** |  |  |  |  |  |  |  |  |
| 0 | 37% | 144 | 44% | 105 | 25% | 39 | 35% | 45 |
| 1 | 44% | 171 | 32% | 77 | 61% | 94 | 46% | 60 |
| 2-4 | 16% | 61 | 18% | 42 | 12% | 19 | 18% | 23 |
| 5+ | 4% | 15 | 6% | 14 | 1% | 1 | 2% | 2 |
| **Vaginal sex partners†** |  |  |  |  |  |  |  |  |
| 0 | 47% | 182 | 55% | 132 | 33% | 50 | 44% | 57 |
| 1 | 39% | 151 | 26% | 63 | 58% | 88 | 40% | 52 |
| 2-4 | 12% | 45 | 14% | 33 | 8% | 12 | 15% | 20 |
| 5+ | 3% | 10 | 4% | 9 | 1% | 1 | 1% | 1 |
| **Oral sex (participant received) partners†** |  |  |  |  |  |  |  |  |
| 0 | 46% | 182 | 48% | 116 | 43% | 66 | 42% | 55 |
| 1 | 36% | 140 | 31% | 74 | 43% | 66 | 40% | 52 |
| 2-4 | 13% | 51 | 13% | 32 | 12% | 19 | 13% | 20 |
| 5+ | 4% | 15 | 6% | 15 | 0% | 0 | 2% | 3 |
| **Oral sex (participant performed) partners†** |  |  |  |  |  |  |  |  |
| 0 | 49% | 199 | 53% | 127 | 47% | 72 | 45% | 58 |
| 1 | 36% | 137 | 30% | 72 | 42% | 65 | 43% | 56 |
| 2-4 | 12% | 41 | 12% | 28 | 8% | 13 | 12% | 16 |
| 5+ | 4% | 11 | 4% | 10 | 1% | 1 | 0% | 0 |
| **Anal sex partners†** |  |  |  |  |  |  |  |  |
| 0 | 88% | 344 | 88% | 212 | 86% | 132 | 89% | 115 |
| 1 | 8% | 32 | 8% | 20 | 8% | 12 | 9% | 12 |
| 2-4 | 3% | 11 | 2% | 6 | 3% | 5 | 1% | 1 |
| 5+ | 0% | 0 | 0% | 0 | 0% | 0 | 0% | 0 |

## Results

### Cohort demographics and potential risk factor prevalence

We recruited 394 participants, 241 in the college-age cohort (ages 18–22 years) and 153 in the older cohort (ages 23+); 130 participants from the full cohort were recruited to also participate in the cervicogenital substudy (Table 1). Approximately two-thirds of our sample was female, and approximately 60% of participants were white. The second most common race was Asian at approximately 20%, though most Asian participants were in the college-age cohort. The majority of participants had never been married, though this varied by age (p<0.001). Approximately 70% of male participants were circumcised. Of the 94% of participants who indicated an HPV vaccination status, approximately half were vaccinated; however, as expected, there were stark differences in vaccination by age, with two-thirds of the college-age cohort reporting vaccination compared to fewer than one quarter of the older cohort (p<0.001). Current alcohol use was similar in the college-age and older cohort, while a larger fraction of the older cohort reported ever smoking cigarettes (p<0.001) and ever using marijuana (p<0.001).

Participants also reported the number of partners in the last 6 months for a variety of intimate and sexual activities (Table 1). Approximately 70% of the full cohort reported at least one recent deep kissing partner, with similar percentages in the different age groups. Fewer participants in the younger cohort reported recent vaginal, oral, or anal sex (56%) compared to the older cohort (74%; p<0.001). Just over half of the full cohort reported at least one recent vaginal sex or oral sex partner, and only 11% reported ever engaging in anal sex. A comparison of cohort characteristic by HPV status is included in the supplement (Table S1).

### HPV prevalence

We found that 10.0% of the 338 participants with a valid oral HPV test at baseline were positive for one or more of the genotypes tested (Table 2). Similarly, 20.0% of the 130 participants’ first valid cervicogenital HPV tests were positive for at least one genotype. When considering only genotypes in the cobas^®^ HPV test, currently the only test FDA-approved for clinical use (on physician-collected cervicogenital swabs), the prevalence of oral HPV was only 6.5%, while the prevalence of cervicogenital HPV was 10.8%.

**Table 2:** HPV prevalence in the oral cavity (valid tests at baseline) or cervicovaginal canal (at first valid test) in the MHOC study by genotype classification (data collected in Ann Arbor, MI, 2015-17, analyzed 2018-2020).

|  | Full cohort<br>(N=338) |  | College-age<br>cohort<br>(N= 241) |  | Older adult<br>cohort<br>(N=153) |  | Cervicogenital<br>substudy cohort<br>(N= 130) |  |
| --- | --- | --- | --- | --- | --- | --- | --- | --- |
|  | % | n | % | n | % | n | % | n |
| Any tested: 6, 11, 16, 18, 31, 35, 33 39, 45, 51, 52, 56, 58, 59, 66, 68, 73, 90 | 10.0% | 34 | 12.6% | 25 | 6.5% | 9 | 20.0% | 26 |
| Clinical test high-risk: 16, 18, 31, 33, 35, 39, 45, 51, 52, 56, 58, 59, 66, 68 | 6.5% | 22 | 8.0% | 16 | 4.3% | 6 | 10.8% | 14 |
| IARC Group 1 and 2A: 16, 18, 33, 31, 35, 39, 45, 51, 52, 56, 58, 59, 68 | 5.3% | 18 | 7.0% | 14 | 2.9% | 4 | 4.6% | 6 |
| Gardasil 9 vaccine genotypes: 6, 11, 16, 18, 31, 33, 45, 52, 58 | 6.5% | 22 | 8.5% | 17 | 3.6% | 5 | 3.8% | 5 |
| Gardasil vaccine genotypes: 6, 11, 16, 18 | 6.2% | 21 | 8.0% | 16 | 3.6% | 5 | 1.5% | 2 |
| Genital warts genotypes: 6, 11 | 2.1% | 7 | 2.5% | 5 | 1.4% | 2 | 1.5% | 2 |

We found oral and cervicogenital HPV prevalence varied by demographic and behavioral variables, though almost no variable achieved significance at level *α*=0.05 in univariable log-binomial regression models (Table 3). Oral HPV prevalence did not differ by sex (10.5% in women vs 9.0% in men); this result held even after age-adjusting the sample. Consistent with the literature, both oral and genital HPV showed a pattern of higher prevalence in younger ages (*<*23) and older ages (50+), with lower prevalence in between. Oral but not cervicogenital HPV was higher in Asian participants, although this result is likely due to the fact that most Asian participants were in the younger cohort. Oral HPV prevalence was not statistically significantly higher in men who were uncircumcised (10.0% vs. 7.6%) and among those who reported ever being diagnosed with an STI (15.8% vs. 9.7%). There were no cervicogenital infections among women ever reporting an STI, but the sample size was small (*n*=9).

**Table 3:** Prevalence of oral and cervicovaginal HPV and prevalence ratios (PR) by covariate in the MHOC study (data collected in Ann Arbor, MI, 2015-17, analyzed 2018-2020). *Other than HPV. **Prevalence ratios are vs “has never engaged in.” †Number of partners in the past 6 months among those who ever engaged in the behavior. ††Cells with fewer than 5 participants are censored.

|  | Oral HPV prevalence<br>(N=338) |  |  |  | Cervicogenital HPV prevalence<br>(N= 130) |  |  |  |
| --- | --- | --- | --- | --- | --- | --- | --- | --- |
|  | % | PR | 95% CI | N | % | OR | 95% CI | N |
| **Age (years)** |  |  |  |  |  |  |  |  |
| 18 | 12.7% | 1 (ref) |  | 79 | 28.6% | 1 (ref) |  | 35 |
| 19-22 | 12.5% | 0.99 | (0.47, 2.09) | 120 | 25.6% | 0.90 | (0.43, 1.86) | 43 |
| 23-29 | 2.3% | 0.18 | (0.02, 1.36) | 44 | 14.3% | 0.50 | (0.13, 2.00) | 14 |
| 30-49 | 6.8% | 0.54 | (0.16, 1.85) | 44 | 0.0% | 0.00 | — | 19 |
| 50+ | 10.0% | 0.77 | (0.43, 1.74) | 51 | 15.8% | 0.55 | (0.17, 1.77) | 19 |
| **Sex** |  |  |  |  |  |  |  |  |
| Female | 10.5% | 1 (ref) |  | 228 | 20.0% | 1 (ref) |  | 130 |
| Male | 9.0% | 0.86 | (0.39, 1.84) | 110 | — | — | — | 0 |
| **Race** |  |  |  |  |  |  |  |  |
| White | 9.6% | 1 (ref) |  | 207 | 20.0% | 1 (ref) |  | 80 |
| Asian | 15.7% | 1.63 | (0.82, 3.22) | 70 | 19.2% | 0.96 | (0.31, 2.37) | 26 |
| Black/Hispanic/multi racial/unknown | 9.1% | 0.94 | (0.24, 3.76) | 22 | 0.0% | 0.00 | — | 10 |
| **Marital status** |  |  |  |  |  |  |  |  |
| Never married | 10.6% | 1 (ref) |  | 255 | 22.7% | 1 (ref) |  | 97 |
| Married/partnered | 6.2% | 0.58 | (0.21, 1.60) | 65 | 16.0% | 0.71 | (0.27, 1.86) | 25 |
| Widowed/divorced/separated | 12.5% | 1.18 | (0.31, 4.53) | 16 | 0.0% | 0.00 | — | 8 |
| **Sexual attraction** |  |  |  |  |  |  |  |  |
| Mostly to only to opposite sex | 11.0% | 1 (ref) |  | 292 | 20.0% | 1 (ref) |  | 120 |
| Equal to only to same sex | 2.7% | 0.25 | (0.03, 1.75) | 37 | 16.7% | 0.83 | (0.13, 5.17) | 6 |
| **Circumcised (male only)** |  |  |  |  |  |  |  |  |
| Yes | 7.6% | 1 (ref) |  | 79 | — | — | — | — |
| No | 10.0% | 1.32 | (0.35, 4.93) | 30 | — | — | — | — |
| **Ever diagnoses with STI\*** |  |  |  |  |  |  |  |  |
| No | 9.7% | 1 (ref) |  | 319 | 21.5% | 1 (ref) |  | 121 |
| Yes | 15.8% | 1.62 | (0.55, 4.84) | 19 | 0.0% | 0.00 | — | 9 |
| **HPV vaccination** |  |  |  |  |  |  |  |  |
| No | 9.4% | 1 (ref) |  | 159 | 18.9% | 1 (ref) |  | 58 |
| Yes | 10.1% | 1.07 | (0.55, 2.08) | 159 | 22.7% | 1.20 | (0.60, 2.40) | 66 |
| **Alcohol use** |  |  |  |  |  |  |  |  |
| Never or non-current | 10.4% | 1 (ref) |  | 96 | 21.6% | 1 (ref) |  | 91 |
| Current | 9.4% | 0.90 | (0.44, 1.83) | 234 | 18.7% | 0.86 | (0.41, 1.83) | 37 |
| **Ever cigarette use** |  |  |  |  |  |  |  |  |
| Never | 10.9% | 1 (ref) |  | 257 | 0.22 | 1 (ref) |  | 103 |
| Ever | 7.5% | 0.69 | (0.30, 1.60) | 80 | 0.12 | 0.52 | (0.17, 1.59) | 26 |
| **Ever marijuana use** |  |  |  |  |  |  |  |  |
| Never | 9.8% | 1 (ref) |  | 150 | 22.1% | 1 (ref) |  | 68 |
| Ever | 8.7% | 0.88 | (0.44, 1.76) | 173 | 18.6% | 0.84 | (0.42, 1.69) | 59 |
| **Has ever engaged in\*\*** |  |  |  |  |  |  |  |  |
| Deep kissing | 8.2% | 0.40 | (0.21, 0.77) | 282 | 18.3% | 0.54 | (0.24, 1.23) | 115 |
| Manual sex | 9.0% | 0.72 | (0.37, 1.42) | 233 | 20.0% | 0.94 | (0.44, 2.04) | 95 |
| Vaginal, oral, or anal sex | 8.9% | 0.69 | (0.35, 1.07) | 257 | 18.9% | 0.78 | (0.36, 1.67) | 101 |
| Vaginal sex | 8.5% | 0.68 | (0.35, 1.30) | 223 | 18.0% | 0.74 | (0.37, 1.48) | 89 |
| Oral sex | 8.6% | 0.65 | (0.33, 1.28) | 243 | 19.4% | 0.88 | (0.41, 1.91) | 98 |
| Anal sex | 9.0% | 0.88 | (0.40, 1.94) | 254 | 15.4% | 0.71 | (0.27, 1.89) | 26 |
| Anilingus | 6.8% | 0.64 | (0.20, 2.00) | 44 | 15.4% | 0.75 | (0.20, 2.81) | 13 |
| **Deep kissing partners†** |  |  |  |  |  |  |  |  |
| 0 | 9.3% | 1 (ref) |  | 43 | 11.1% | 1 (ref) |  | 18 |
| 1 | 7.8% | 0.84 | (0.28, 2.50) | 141 | 9.3% | 0.83 | (0.18, 3.93) | 54 |
| 2-4 | 6.3% | 0.68 | (0.18, 2.58) | 63 | 29.0% | 2.6 | (0.63, 10.8) | 31 |
| 5+ | 11.4% | 1.23 | (0.33, 4.56) | 35 | 41.7% | 3.75 | (0.86, 16.3) | 12 |
| **Vaginal, oral, or anal sex partners†** |  |  |  |  |  |  |  |  |
| 0 | 14.3% | 1 (ref) |  | 119 | 15.6% | 1 (ref) |  | 45 |
| 1 | 6.7% | 0.47 | (0.22, 0.98) | 150 | 15.0% | 0.96 | (0.39, 2.39) | 60 |
| 2-4 | 7.5% | 0.53 | (0.19, 1.49) | 53 | 39.1% | 2.52 | (1.07, 5.88) | 23 |
| 5+ | 15.4% | 1.08 | (0.28, 4.14) | 13 | †† | †† | †† | 2 |
| **Vaginal sex partners<sup>†</sup>** |  |  |  |  |  |  |  |  |
| 0 | 13.5% | 1 (ref) |  | 148 | 17.5% | 1 (ref) |  | 57 |
| 1 | 5.1% | 0.38 | (0.16, 0.86) | 138 | 13.4% | 0.77 | (0.32, 1.86) | 52 |
| 2-4 | 7.9% | 0.58 | (0.18, 1.86) | 38 | 45.0% | 2.57 | (1.22, 5.39) | 20 |
| 5+ | 33.3% | 2.47 | (0.90, 6.77) | 9 | †† | †† | †† | 1 |
| **Oral sex (participant received) partners<sup>†</sup>** |  |  |  |  |  |  |  |  |
| 0 | 12.9% | 1 (ref) |  | 155 | 16.4% | 1 (ref) |  | 55 |
| 1 | 5.9% | 0.46 | (0.20, 1.05) | 118 | 19.2% | 1.18 | (0.52, 2.66) | 52 |
| 2-4 | 10.6% | 0.82 | (0.33, 2.08) | 47 | 30.0% | 1.83 | (0.75, 4.50) | 20 |
| 5+ | 7.7% | 0.60 | (0.09, 4.09) | 13 | †† | †† | †† | 3 |
| **Oral sex (participant performed) partners<sup>†</sup>** |  |  |  |  |  |  |  |  |
| 0 | 13.1% | 1 (ref) |  | 168 | 15.5% | 1 (ref) |  | 58 |
| 1 | 5.0% | 0.38 | (0.16, 0.91) | 121 | 17.9% | 1.15 | (0.51, 2.62) | 56 |
| 2-4 | 14.3% | 1.09 | (0.44, 2.68) | 35 | 43.8% | 2.82 | (1.24, 6.39) | 16 |
| 5+ | 0.0% | 0.00 | — | 9 | †† | †† | †† | 0 |
| **Anal sex partners<sup>†</sup>** |  |  |  |  |  |  |  |  |
| 0 | 9.9% | 1 (ref) |  | 294 | 20.0% | 1 (ref) |  | 115 |
| 1 | 7.4% | 0.75 | (0.19, 2.98) | 27 | 25.0% | 1.25 | (0.44, 3.55) | 12 |
| 2-4 | 18.2% | 1.84 | (0.50, 6.77) | 11 | †† | †† | †† | 1 |

There was little difference in oral or cervicogenital prevalence among those who did and did not report vaccination, even when considering only genotypes 6, 11, 16, and 18 (oral prevalence among vaccinated (6.3%) and unvaccinated (6.3%); cervicogenital prevalence among vaccinated (1.5%) and unvaccinated (1.7%)) although the absolute numbers of positives are small. The apparent lack of protection from vaccination appears to be the result of an ecological fallacy, however. When considering the college-age and older adult cohort separately, oral HPV prevalence is lower in vaccinated individuals of both groups (Table 4).

**Table 4:** Prevalence of oral HPV and prevalence ratios (PR) by covariate in the MHOC study in the college-age and older adult cohorts (data collected in Ann Arbor, MI, 2015-17, analyzed 2018-2020). *Other than HPV. **Prevalence ratios are vs “has never engaged in.” †Number of partners in the past 6 months among those who ever engaged in the behavior. ††Cells with fewer than 5 participants are censored.

|  | Oral HPV prevalence in the college-age cohort (N=199) |  |  |  | Oral HPV prevalence in the older adult cohort (N=139) |  |  |  |
| --- | --- | --- | --- | --- | --- | --- | --- | --- |
|  | % | OR | 95% CI | N | % | OR | 95% CI | N |
| **Age** |  |  |  |  |  |  |  |  |
| 18 | 12.7% | 1 (ref) |  | 79 | — | — | — | 0 |
| 19-22 | 12.5% | 0.99 | (0.47, 2.09) | 120 | — | — | — | 0 |
| 23-29 | — | — | — | 0 | 2.3% | 1 (ref) |  | 44 |
| 30-49 | — | — | — | 0 | 6.8% | 3.00 | (0.32, 27.7) | 44 |
| 50+ | — | — | — | 0 | 9.8% | 4.31 | (0.52, 35.5) | 51 |
| **Sex** |  |  |  |  |  |  |  |  |
| Female | 13.7% | 1 (ref) |  | 139 | 5.6% | 1 (ref) |  | 89 |
| Male | 10.0% | 0.73 | (0.31, 1.74) | 60 | 8.0% | 1.42 | (0.40, 5.06) | 50 |
| **Race** |  |  |  |  |  |  |  |  |
| White | 12.0% | 1 (ref) |  | 108 | 7.1% | 1 (ref) |  | 99 |
| Asian | 16.4% | 1.36 | (0.64, 2.92) | 61 | 11.1% | 1.57 | (0.22, 11.4) | 9 |
| Black/Hispanic/multi racial/unknown | 18.1% | 1.51 | (0.39, 5.85) | 11 | 0.0% | — | — | 11 |
| **Marital status** |  |  |  |  |  |  |  |  |
| Never married | 12.2% | 1 (ref) |  | 196 | 5.1% | 1 (ref) |  | 59 |
| Married/partnered | †† | †† | †† | 1 | 6.3% | 1.23 | (0.29, 5.26) | 64 |
| Widowed/divorced/separated | †† | †† | †† | 1 | 13.3% | 2.62 | (0.48, 14.3) | 15 |
| **Sexual attraction** |  |  |  |  |  |  |  |  |
| Mostly to only to opposite sex | 14.2% | 1 (ref) |  | 169 | 6.5% | 1 (ref) |  | 123 |
| Equal to only to same sex | 0.0% | — | — | 24 | 7.7% | 1.18 | (0.16, 8.73) | 13 |
| **Circumcised (male only)** |  |  |  |  |  |  |  |  |
| Yes | 7.5% | 1 (ref) |  | 40 | 7.7% | 1 (ref) |  | 39 |
| No | 10.5% | 1.40 | (0.26, 7.71) | 19 | 9.1% | 1.18 | (0.14, 10.3) | 11 |
| **Ever diagnoses with STI\*** |  |  |  |  |  |  |  |  |
| No | 12.8% | 1 (ref) |  | 196 | 4.9% | 1 (ref) |  | 123 |
| Yes | †† | †† | †† | 3 | 18.8% | 3.84 | (1.06, 13.9) | 16 |
| **HPV vaccination** |  |  |  |  |  |  |  |  |
| No | 14.0% | 1 (ref) |  | 50 | 7.34% | 1 (ref) |  | 109 |
| Yes | 11.6% | 0.83 | (0.36, 1.92) | 129 | 3.33% | 0.45 | (0.06, 3.49) | 30 |
| **Alcohol use** |  |  |  |  |  |  |  |  |
| Never or non-current | 14.8% | 1 (ref) |  | 61 | 2.9% | 1 (ref) |  | 35 |
| Current | 10.8% | 0.73 | (0.33, 1.59) | 130 | 7.7% | 2.69 | (0.35, 20.8) | 104 |
| **Ever cigarette use** |  |  |  |  |  |  |  |  |
| Never | 13.1% | 1 (ref) |  | 175 | 6.1% | 1 (ref) |  | 82 |
| Ever | 8.3% | 0.63 | (0.16, 2.52) | 24 | 7.1% | 1.17 | (0.33, 4.17) | 56 |
| **Ever marijuana use** |  |  |  |  |  |  |  |  |
| Never | 13.9% | 1 (ref) |  | 115 | 1.7% | 1 (ref) |  | 75 |
| Ever | 9.3% | 0.67 | (0.29, 1.55) | 75 | 8.0% | 4.64 | (0.57, 37.5) | 58 |
| **Has ever engaged in\*\*** |  |  |  |  |  |  |  |  |
| Deep kissing | 10.1% | 0.44 | (0.21, 0.91) | 159 | 5.7% | 0.43 | (0.10, 1.87) | 123 |
| Manual sex | 10.8% | 0.74 | (0.35, 1.56) | 120 | 7.1% | 1.56 | (0.20, 11.8) | 113 |
| Vaginal, oral, or anal sex | 11.1% | 0.78 | (0.36, 1.66) | 126 | 6.9% | — | — | 131 |
| Vaginal sex | 10.2% | 0.71 | (0.33, 1.53) | 98 | 7.2% | — | — | 125 |
| Oral sex | 9.9% | 0.62 | (0.29, 1.31) | 121 | 7.4% | — | — | 122 |
| Anal sex | 12.0% | 0.97 | (0.31, 3.03) | 25 | 7.5% | 1.25 | (0.35, 4.46) | 53 |
| Anilingus | 7.1% | 0.55 | (0.08, 3.75) | 14 | 6.7% | 1.02 | (0.22, 4.65) | 30 |
| **Deep kissing partners†** |  |  |  |  |  |  |  |  |
| 0 | 5.9% | 1 (ref) |  | 17 | 11.5% | 1 (ref) |  | 26 |
| 1 | 13.1% | 2.23 | (0.30, 16.6) | 61 | 3.8% | 0.33 | (0.07, 1.51) | 80 |
| 2-4 | 8.3% | 1.42 | (0.17, 11.8) | 48 | 0.0% | — | — | 15 |
| 5+ | 9.1% | 1.54 | (0.17, 13.8) | 33 | †† | †† | †† | 2 |
| **Vaginal, oral, or anal sex partners†** |  |  |  |  |  |  |  |  |
| 0 | 15.5% | 1 (ref) |  | 84 | 11.4% | 1 (ref) |  | 35 |
| 1 | 9.5% | 0.62 | (0.25, 1.53) | 63 | 4.6% | 0.40 | (0.11, 1.52) | 87 |
| 2-4 | 8.1% | 0.52 | (0.16, 1.72) | 37 | 6.3% | 0.55 | (0.07, 4.51) | 16 |
| 5+ | 21.4% | 1.38 | (0.45, 4.25) | 14 | — | — | — | 0 |
| **Vaginal sex partners<sup>†</sup>** |  |  |  |  |  |  |  |  |
| 0 | 15.4% | 1 (ref) |  | 104 | 9.1% | 1 (ref) |  | 44 |
| 1 | 7.3% | 0.47 | (0.17, 1.35) | 55 | 3.6% | 0.40 | (0.09, 1.70) | 83 |
| 2-4 | 7.1% | 0.46 | (0.11, 1.90) | 28 | 10.0% | 1.10 | (0.14, 8.81) | 10 |
| 5+ | 25.0% | 1.63 | (0.45, 5.86) | 8 | † | † | † | 1 |
| **Oral sex (participant received) partners<sup>†</sup>** |  |  |  |  |  |  |  |  |
| 0 | 15.8% | 1 (ref) |  | 95 | 8.3% | 1 (ref) |  | 60 |
| 1 | 8.6% | 0.55 | (0.21, 1.42) | 58 | 3.3% | 0.40 | (0.08, 1.98) | 60 |
| 2-4 | 10.3% | 0.66 | (0.20, 2.11) | 29 | 11.1% | 1.33 | (0.49, 6.30) | 18 |
| 5+ | 7.7% | 0.49 | (0.07, 3.39) | 13 | — | — | — | 0 |
| **Oral sex (participant performed) partners<sup>†</sup>** |  |  |  |  |  |  |  |  |
| 0 | 16.3% | 1 (ref) |  | 104 | 7.8% | 1 (ref) |  | 64 |
| 1 | 6.9% | 0.42 | (0.15, 1.19) | 58 | 3.2% | 0.41 | (0.08, 2.01) | 63 |
| 2-4 | 12.0% | 0.73 | (0.23, 2.31) | 25 | 20.0% | 2.56 | (0.57, 11.5) | 10 |
| 5+ | 0.0% | — | — | 8 | †† | †† | †† | 1 |
| **Anal sex partners<sup>†</sup>** |  |  |  |  |  |  |  |  |
| 0 | 12.7% | 1 (ref) |  | 173 | 5.8% | 1 (ref) |  | 121 |
| 1 | 5.9% | 0.46 | (0.07, 3.22) | 17 | 10.0% | 1.63 | (0.24, 12.7) | 10 |
| 2-4 | 16.7% | 1.31 | (0.21, 8.18) | 6 | 20.0% | 3.46 | (0.52, 23.0) | 5 |

Based on participant self-reporting of sexual partners, we found a high prevalence of oral and cervicogenital HPV infection among those resulted in prevalence ratio point estimates for those reporting deep kissing (0.4, 95% CI: 0.21, 0.77) or any sexual behavior (not statistically significant) that were less than 1 (Table 3). This directionality was consistent across the college-age and older adult cohorts (Table 4). Among those reporting at least one recent sexual partner for each behavior type, point estimates of prevalence increased with the number of sexual partners for both oral and cervicogenital infection (Table 3), with few exceptions, though only a few prevalence ratios reach statistical significance (including, for cervicogenital HPV infection, 2-4 vaginal, oral, or anal sex partners; 2-4 vaginal sex partners; and 2-4 sex partners where the participant performed oral sex).

## Discussion

This analysis presents a first look at the Michigan HPV and Oropharyngeal Cancer (MHOC) Study’s epidemiological cohort. In this baseline analysis, we investigated associations of HPV prevalence with demographic and behavioral risk factors. Compared to the nationally representative National Health and Nutrition Examination Survey [10, 11], we find much higher oral HPV prevalence among women (NHANES 2009–14: 3.4%; MHOC: 10.5%), especially considering that NHANES tests for 37 genotypes while MHOC tested for 18. For IARC Group 1 HPV types [22], oral prevalence in women in NHANES 2009–14 was 1.3%, compared to 6.0% in MHOC. For men, Group 1 oral HPV prevalence in NHANES was 5.6% (11.1% for any tested genotype) and in MHOC was 4.8% (9.0% for any tested). Interestingly, the high oral HPV prevalence in women is not matched by a high cervicogenital prevalence; Group 1 cervicogenital prevalence in NHANES was 18.6% (40.6% for any tested genotype) and in MHOC was 4.6% (20.0% for any tested). In MHOC, oral HPV prevalence in women was higher than men in the younger cohort (13.7% vs. 10.0%) but lower in the older cohort (5.6% vs 8.0%). Although the MHOC study was not designed to be nationally representative, these differences persist after age-adjusting the prevalence. Given the similarity in prevalence between NHANES and MHOC among men, the differences between the two studies are unlikely to be purely based on assay sensitivity or laboratory methods. One explanation for the high HPV prevalence in women in MHOC is that prevalence of HPV—both overall prevalence and genotype-specific prevalence—may vary regionally. It is also possible that the risk behaviors of MHOC participants differ from the national average in ways that make oral infection more likely and cervicogenital HPV less likely.

However, while the prevalence magnitudes differ between NHANES and MHOC, the age-specific patterns are very similar, both exhibiting the classic bimodal distribution [26]: high prevalence among the youngest and oldest age groups, with lower prevalence in between. This underscores the importance of examining age and cohort patterns in HPV prevalence, as the increase in higher age groups may be the result of generational (birth-cohort) differences, increased sexual activity at older ages, or reactivation of HPV at older ages [10, 27–29].

We find high oral and cervicogenital prevalence among people who report no recent sexual partners. This result is not simply an age effect (as might be the case if younger people were less likely to have sexual partners but had a higher prevalence overall) as it persists when we separate the younger and older cohorts. While it is possible that some infections may persist from previous sexual partners (approximately 1/3 of people with no recent sexual partners reported at least one lifetime sexual partners), it may be more likely that our participants are not reporting their experiences accurately, possibly to avoid consequences if their answers were ever disclosed or simply to minimize the survey burden. In any case, we believe that our analysis of the associations of sexual behavior and HPV prevalence are subject to some level of misclassification bias. However, if considering only those who report at least one recent sexual partner, prevalence increases with the number of recent sex partners, as expected. While sexual acts are strongly correlated, previous studies have differed in the reported strength of association between oral vs. vaginal sex and oral HPV prevalence [12–15]; here, we find that both oral and cervicogenital HPV prevalence are strongly associated with number of recent vaginal sex partners (among those reporting ever having had vaginal sex). The number of oral sex partners (among those reporting ever having had oral sex), while still positively associated with both oral and cervicogenital HPV prevalence, was not substantially stronger than the number of recent deep kissing partners (among those reporting ever deep kissing). Although there is reason to believe that risk differs based on the sex of one’s partner [16], we were not powered to investigate these differences in this study.

Nationally representative data demonstrate that oral and genital prevalence of HPV vaccine genotypes is reduced among vaccinated people [10, 30]. In this analysis, we were not able to find strong evidence of a reduction in prevalence likely due to not having a large enough sample size. In addition, the lack of difference at the cohort level is, in part, an ecological fallacy, as vaccination is protective when considering the younger and older cohorts separately. Misreporting of vaccination status likely accounts for the smaller than expected observed effect of vaccination in this analysis. Vaccination is also known to be less effective in people already exposed to HPV [31], which may further attenuate effects here.

The limitations of this baseline analysis include the relatively small sample size compared to NHANES and other HPV prevalence studies. However, because this cohort is designed to be studied longitudinally, we will have substantially more power in the analysis of HPV prevalence over time. Analysis of the longitudinal data will later provide a detailed consideration of the short-term dynamics of the detection of oral and cervicogenital HPV [32]. This in turn will aid in the development of oral HPV and sexual behavior transmission models, and models of the natural history of oropharyngeal cancer [21, 33]. Other limitations of the study are the self-reported nature of sexual and other behaviors and vaccination, which are subject to misclassification.

This hypothesis-generating work contributes to our understanding of the prevalence and determinants of oral HPV by suggesting that prevalence of HPV in women is not uniformly lower than in men. More work is needed to understand regional variation in oral HPV prevalence overall and by genotype. Too, more work is needed to better understand the specific transmission pathways that lead to oral HPV infection, particularly the relative contribution of oral sex vs autoinoculation from genital infection and the role of latent infections.

## Data Availability

The datasets generated and/or analyzed during the current study are not publicly available because of participant privacy concerns but are available from the corresponding author on reasonable request. IRB approval or a data use agreement may be required.

## Acknowledgments

This work was supported by National Institutes for Health grant U01CA182915. Data management was supported by the Michigan Institute for Clinical & Health Research (CTSA grant UL1TR002240). We would like to thank the Michigan HPV and Oropharyngeal Cancer (M-HOC) study team for making this work possible, including Renata Terrazzan, Eliyas Asfaw, Jung Woo Lee, Alexandra Kalabat, Ivan Montoya, Courtney Walsh, Ashley Wu, Liana Ysabel Bautista, Anna Morris, Nadine Jawad, Manila Hada, Bala Naveen Kakaraparthi, Peter Tortora, Taylor Vandenberg, Christina Hanson, Lucy Yang, Macy Afsari, Alanna Clark, Anna Gottschlich, Chinmay Pandit, Greg Foakes, Kristin Bevilacqua, Jesse Contreras, Maxwell Salvatore, Christian Alvarez, Pianpian Cao, Kelly Sun, Sheila Terrones, Lisa Petersen, and Miranda West.

## References

[1] Jemal A, Simard EP, Dorell C, Noone AM, Markowitz LE, Kohler B, et al. Annual Report to the Nation on the Status of Cancer, 1975-2009, Featuring the Burden and Trends in Human Papillomavirus (HPV)-Associated Cancers and HPV Vaccination Coverage Levels. Journal of the National Cancer Institute. 2013;105(3):175–201.

[2] Gillison ML, Alemany L, Snijders PJF, Chaturvedi A, Steinberg BM, Schwartz S, et al. Human papillomavirus and diseases of the upper airway: head and neck cancer and respiratory papillomatosis. Vaccine. 2012;30 Suppl 5:F34–54.

[3] Serrano B, Brotons M, Bosch FX, Bruni L. Epidemiology and burden of HPV-related disease. Best Practice & Research Clinical Obstetrics & Gynaecology. 2018;47:14–26.

[4] de Martel C, Plummer M, Vignat J, Franceschi S. Worldwide burden of cancer attributable to HPV by site, country and HPV type. International journal of cancer. 2017;141(4):664–670.

[5] Chaturvedi AK, Engels EA, Anderson WF, Gillison ML. Incidence trends for human papillomavirus-related and -unrelated oral squamous cell carcinomas in the United States. Journal of Clinical Oncology. 2008;26(4):612–9.

[6] Chaturvedi AK, Engels EA, Pfeiffer RM, Hernandez BY, Xiao W, Kim E, et al. Human papillomavirus and rising oropharyngeal cancer incidence in the United States. Journal of Clinical Oncology. 2011;29(32):4294–301.

[7] Chaturvedi AK, Anderson WF, Lortet-Tieulent J, Paula Curado M, Ferlay J, Franceschi S, et al. Worldwide trends in incidence rates for oral cavity and oropharyngeal cancers. Journal of Clinical Oncology. 2013;31(36):4550–4559.

[8] Chaturvedi AK, D’Souza G, Gillison ML, Katki HA. Burden of HPV-positive oropharynx cancers among ever and never smokers in the U.S. population. Oral Oncology. 2016;60:61–67.

[9] Walline HM, Komarck C, McHugh JB, Byrd SA, Spector ME, Hauff SJ, et al. High-risk human papillomavirus detection in oropharyngeal, nasopharyngeal, and oral cavity cancers comparison of multiple methods. JAMA Otolaryngology - Head and Neck Surgery. 2013;139(12):1320–7.

[10] Brouwer AF, Eisenberg MC, Carey TE, Meza R. Multisite HPV infections in the United States (NHANES 20032014): An overview and synthesis. Preventive Medicine. 2019;123:288–298.

[11] Chaturvedi AK, Graubard BI, Broutian T, Xiao W, Pickard RKL, Kahle L, et al. Prevalence of Oral HPV Infection in Unvaccinated Men and Women in the United States, 2009-2016. JAMA. 2019;322(10):977.

[12] Edelstein ZR, Schwartz SM, Hawes S, Hughes JP, Feng Q, Stern ME, et al. Rates and determinants of oral human papillomavirus infection in young men. Sexually Transmitted Diseases. 2012;39(11):860–7.

[13] Cook RL, Thompson EL, Kelso NE, Friary J, Hosford J, Barkley P, et al. Sexual Behaviors and Other Risk Factors for Oral Human Papillomavirus Infections in Young Women. Sexually Transmitted Diseases. 2014;41(8):486–492.

[14] The prevalence and incidence of oral human papillomavirus infection among young men and women, aged 18-30 years. Sexually Transmitted Diseases. 2012;39(7):559–66.

[15] D’Souza G, Cullen K, Bowie J, Thorpe R, Fakhry C. Differences in oral sexual behaviors by gender, age, and race explain observed differences in prevalence of oral human papillomavirus infection. PLOS One. 2014;9(1):e86023.

[16] D’Souza G, Wentz A, Kluz N, Zhang Y, Sugar E, Youngfellow RM, et al. Sex Differences in Risk Factors and Natural History of Oral Human Papillomavirus Infection. Journal of Infectious Diseases. 2016;213(12):1893–1896.

[17] Chung CH, Bagheri A, D’Souza G. Epidemiology of oral human papillomavirus infection. Oral Oncology. 2014;50(5):364–369.

[18] Chaturvedi AK, Graubard BI, Broutian T, Pickard RKL, Tong ZY, Xiao W, et al. Effect of prophylactic human papillomavirus (HPV) vaccination on oral HPV infections among young adults in the United States. Journal of Clinical Oncology. 2018;36(3):262–267.

[19] Schlecht NF, Masika M, Diaz A, Nucci-Sack A, Salandy A, Pickering S, et al. Risk of Oral Human Papillomavirus Infection Among Sexually Active Female Adolescents Receiving the Quadrivalent Vaccine. JAMA Network Open. 2019;2(10):e1914031.

[20] Brouwer AF, Meza R, Eisenberg MC. Transmission heterogeneity and autoinoculation in a multisite infection model of HPV. Mathematical Biosciences. 2015 dec;270:115–125.

[21] Eisenberg MC, Campredon LP, Brouwer AF, Walline HM, Marinelli BM, Lau YK, et al. Dynamics and Determinants of HPV Infection: The Michigan HPV and Oropharyngeal Cancer (M-HOC) Study. BMJ Open. 2018;8(10):e021618.

[22] International Agency for Research on Cancer. Biological Agents; 2012.

[23] Harris PA, Taylor R, Thielke R, Payne J, Gonzalez N, Conde JG. Research electronic data capture (REDCap)a metadata-driven methodology and workflow process for providing translational research informatics support. Journal of Biomedical Informatics. 2009;42(2):377–381.

[24] Harris PA, Taylor R, Minor BL, Elliott V, Fernandez M, O’Neal L, et al. The REDCap consortium: Building an international community of software platform partners. Journal of Biomedical Informatics. 2019;95:103208.

[25] R Core Team. R: A Language and Environment for Statistical Computing. Vienna, Austria; 2020. Available from: https://www.R-project.org/.

[26] Gillison ML, Broutian T, Pickard RKL, Tong ZyZy, Xiao W, Kahle L, et al. Prevalence of oral HPV infection in the United States, 2009-2010. Journal of the American Medical Association. 2012;307(7):693–703.

[27] Chaturvedi AK, Graubard BI, Pickard RKL, Xiao W, Gillison ML. High-Risk Oral Human Papillomavirus Load in the US Population, National Health and Nutrition Examination Survey 20092010. Journal of Infectious Diseases. 2014;210:441–447.

[28] Gravitt PE, Rositch AF, Silver MI, Marks MA, Chang K, Burke AE, et al. A cohort effect of the sexual revolution may be masking an increase in human papillomavirus detection at menopause in the United States. Journal of Infectious Diseases. 2013;207(2):272–280.

[29] Paul P, Hammer A, Rositch AF, Burke AE, Viscidi RP, Silver MI, et al. Rates of New Human Papillomavirus Detection and Loss of Detection in Middle-aged Women by Recent and Past Sexual Behavior. The Journal of Infectious Diseases. 2021;223(8):1423–1432.

[30] Chaturvedi AK, Graubard BI, Broutian T, Pickard RK, Tong ZY, Xiao W, et al. Effect of prophylactic human papillomavirus (HPV) vaccination on oral HPV infections among young adults in the United States. Journal of Clinical Oncology. 2018;36(3):262.

[31] Harper DM, Paavonen J. Age for HPV vaccination. Vaccine. 2008;26:A7–A11.

[32] Brouwer AF, Campredon LP, Walline HM, Marinelli BM, Goudsmit CM, Thomas TB, Delinger RL, Lau YK, Andrus EC, Nair T, Carey TE, Eisenberg MC, Meza R. Incidence and clearance of oral and cervicogenital HPV infection: longitudinal analysis of the MHOC cohort study. BMJ Open. 2022;12:1–9

[33] Brouwer AF, Eisenberg MC, Meza R. Case Studies of Gastric, Lung, and Oral Cancer Connect Etiologic Agent Prevalence to Cancer Incidence. Cancer Research. 2018;78(12):3386–3397.

