## Supplementary material for "Prevalence and determinants of oral and cervicogenital HPV infection: baseline analysis of the MHOC cohort study": Table S1

### Supplement

Table S1: Baseline characteristics of participants in the MHOC Study by oral HPV status (data collected in Ann Arbor, MI, 2015-17, analyzed 2018-2020). Note: percentages may not add up to 100% as participants could refuse to answer questions. *Other than HPV.

| | Oral HPV- (N=304) | | Oral HPV+ (N=34) | |
|---|---|---|---|---|
| | % | n | % | n |
| **Age** | | | | |
| 18 | 23% | 69 | 29% | 10 |
| 19-22 | 35% | 105 | 44% | 15 |
| 23-29 | 14% | 43 | 3% | 1 |
| 30-49 | 13% | 41 | 9% | 3 |
| 50+ | 15% | 46 | 15% | 5 |
| **Sex** | | | | |
| Female | 67% | 204 | 71% | 24 |
| Male | 33% | 100 | 29% | 10 |
| **Race** | | | | |
| White | 62% | 187 | 59% | 11 |
| Asian | 19% | 59 | 32% | 20 |
| Black/Hispanic/multiracial/unknown | 19% | 58 | 9% | 3 |
| **Marital status** | | | | |
| Never married | 75% | 228 | 79% | 27 |
| Married/partnered | 20% | 61 | 12% | 4 |
| Widowed/divorced/separated | 5% | 14 | 6% | 2 |
| **Sexual attraction** | | | | |
| Mostly to only to opposite sex | 86% | 260 | 94% | 32 |
| Equal to only to same sex | 12% | 36 | 3% | 1 |
| **Circumcised (male only)** | | | | |
| Yes | 73% | 73 | 60% | 6 |
| No | 27% | 27 | 30% | 3 |
| **Ever diagnoses with STI*** | | | | |
| No | 95% | 228 | 91% | 31 |
| Yes | 5% | 16 | 9% | 3 |
| **HPV vaccination** | | | | |
| No | 47% | 144 | 44% | 15 |

| | | | | |
|---|---|---|---|---|
| Yes | 47% | 143 | 47% | 16 |
| **Alcohol use** | | | | |
| Never or non-current | 28% | 86 | 29% | 10 |
| Current | 70% | 212 | 65% | 22 |
| **Ever cigarette use** | | | | |
| Never | 75% | 229 | 82% | 28 |
| Ever | 24% | 74 | 18% | 6 |
| **Ever marijuana use** | | | | |
| Never | 51% | 156 | 50% | 17 |
| Ever | 45% | 137 | 38% | 13 |
| **Has ever engaged in** | | | | |
| Deep kissing | 85% | 259 | 67% | 23 |
| Manual sex | 70% | 212 | 62% | 21 |
| Vaginal, oral, or anal sex | 77% | 234 | 67% | 23 |
| Vaginal sex | 67% | 204 | 56% | 19 |
| Oral sex | 73% | 222 | 62% | 21 |
| Anal sex | 23% | 71 | 21% | 7 |
| Anilingus | 13% | 41 | 9% | 3 |
